# Distinct Retrotransposon Transcriptome in Pediatric Crohn’s Disease

**DOI:** 10.1101/2025.04.21.25326010

**Authors:** Qing Chen, Colton McNinch, Eve May, Phoebe LaPoint, Galina Koroleva, Ashleigh Sutton, Ruhika Prasad, Diana Jo, Brynn O’Laughlin, Anal Patel, Shira Levy, Suchitra Hourigan

**Affiliations:** Clinical Microbiome Unit, Laboratory of Host Immunity and Microbiome, Division of Intramural Research National Institute of Allergy and Infectious Diseases, National Institutes of Health, Bethesda, Maryland, United States; Bioinformatics and Computational Biosciences Branch National Institute of Allergy and Infectious Diseases, National Institutes of Health, Bethesda, Maryland, United States; NIH Center for Human Immunology, National Institutes of Health, Bethesda, Maryland, United States

**Keywords:** inflammatory bowel disease, long interspersed nuclear element-1, human endogenous retroviruses, gene expression

## Abstract

**Background and Aims:** Crohn’s disease (CD) is an autoimmune condition with inflammation of the gastrointestinal tract. The etiology of CD is complex with underlying mechanisms only partially elucidated. Retrotransposons, a category of transposable elements within a genome, have been shown to contribute to the pathogenesis of inflammatory and autoimmune disorders. This research aimed to explore the expression of retrotransposons in children with CD.

**Methods:** To assess the expression level of retrotransposons, high-throughput expression analysis at the locus level was performed including LINE, SINE, and LTR-retrotransposons (human endogenous retroviruses (HERVs)) in total RNAseq of ileal and rectal biopsies from treatment-naïve children with CD and age-matched non-Inflammatory Bowel Disease (IBD) controls (CMU-dataset). Findings were validated in public datasets (GSE57945 Ileal; GSE117993 Rectal) from the pediatric RISK study.

**Results:** Consistent separation of retrotransposon expression between CD and non-IBD controls in both the CMU-dataset and the RISK-dataset was observed in ileal biopsies, but less so in rectal biopsies. In total, 118 differentially expressed retrotransposon loci were identified (27 upregulated and 91 downregulated; 74 LINE-1 and 44 HERV) in CMU-ileal dataset. Fifteen retrotransposon loci were consistently downregulated in both datasets (CMU-ileal and GSE57945): HERV9N-int, HERV9-int (2 loci), HERVH-int (2 loci), HERVK22-int, LTR5A, HERVK-int, L1PA4 (5 loci), L1PA6, L1PA14.

**Conclusions:** Retrotransposon transcriptome in children with CD significantly differed from non-IBD controls in ileal biopsies with 15 retrotransposon loci consistently downregulated in CD. The possible regulatory function of these retrotransposon loci merits additional research. This study may provide valuable perspectives for the advancement of novel therapeutic strategies.

## Introduction

Crohn’s disease (CD), an inflammatory bowel disease (IBD) affecting around 200/100,000 people, causes significant morbidity, reduces quality of life, and represents a substantial healthcare burden ^1,2^. While CD is multifactorial in etiology, including genetic predisposition and gut microbiome imbalance, its pathogenesis remains incompletely understood. Although effective treatments (not cures) exist, more therapeutic targets are needed to develop new therapies, as there is often a loss of response over time to current treatments ^3^.

Retrotransposons, a type of transposable element that can successfully expand while retaining transcriptional activity, are highly repetitive strands of DNA that are mobilized by an RNA intermediate in the human genome ^4^. Retrotransposons make up around 40% of the human genome and include LINE (long interspersed nuclear elements), SINE (short interspersed nuclear elements), and LTR-retrotransposons, which are also known as human endogenous retroviruses (HERVs) ^5^. The activity of retrotransposons affects the human genome in many ways, including their potential to modulate pathologic genes and cause molecular mimicry, contributing to cancer and autoimmune/inflammatory diseases ^4,6-8^. Notably, adults with CD showed lower LINE-1 methylation without expression level changes in intestinal mucosa ^9^ and altered HERV expression in biopsies and blood compared to controls^10-12^.

Although evidence suggests involvement of LINE-1 and HERV in CD, a comprehensive locus-level assessment of the retrotransposon transcriptome in CD is lacking. Moreover, this has not been investigated in children, whose shorter environmental exposure minimizing age-related changes in retrotransposon expression may enable identification of key retrotransposons in CD development ^13^. Therefore, the retrotransposon transcriptome was investigated in ileal and rectal biopsies from a prospective cohort of treatment-naïve children with CD and age-matched controls by analyzing total RNA seq datasets with technical parameters allowing for accurate read mapping to highly repetitive genomic regions. Findings were further validated in public datasets from the RISK Cohort ^14,15^.

## Methods

### Cohort

Children undergoing colonoscopy were enrolled with written informed consent/assent (Inova protocol ID: U21-02-4394). Children were either diagnosed with CD (n=8) or had no eventual diagnosis of gut inflammation and were considered as non-IBD controls (n=12), **Table 1**. Ileal and rectal biopsies were obtained and stored in RNAlater at −80°C until analysis. Public datasets were from the RISK study: GSE57945 for ileal (CD = 204, non-IBD controls = 42) and GSE117993 for rectal (CD = 36, non-IBD controls = 20) ^14,15^.

**Table 1:** Demographic characteristics of CMU-cohort.

|  | **Non-IBD control (n=12)** | **CD (n=8)** |
| --- | --- | --- |
| Age in years, mean (range) | 14 (7-22) | 13 (4-16) |
| Male (n) | 5 | 6 |
| Female (n) | 7 | 2 |
| Ileal biopsy (n) | 11 | 5 |
| Rectal biopsy (n) | 12 | 8 |
| Disease location (n) |  | Terminal ileum (3)<br>Colon (3)<br>Terminal ileum and colon (2) |
| Race (n) | White (8)<br>African American (2)<br>Native Hawaiian (1)<br>More than one race (1) | White (6)<br>Native Alaska (1)<br>Asian (1) |
| Ethnicity (n) | Hispanic or Latino (1) | Hispanic or Latino (0) |

### RNA-sequencing

Total RNA was extracted using the AllPrep DNA/RNA Micro Kit (QIAGEN). Libraries were prepared using the Universal Plus Total RNA-seq kit (TECAN) and sequenced on the Illumina NextSeq platform with paired-end (2 x 151bp).

### Statistical analysis

Retrotransposon and gene expression analyses were conducted using the *ervx* ^16^and RNA-Seek pipelines ^17^, respectively. Both pipelines began with adapter removal and quality trimming using Cutadapt v1.18 ^18^. Cleaned reads were aligned to the human genome (GENCODE v20 primary assembly, GRCh38) using STAR v2.7.6a in two-pass basic mode ^19^. Retrotransposon expression was quantified across all human loci cataloged in the Genome-based Endogenous Viral Element Database ^20^ using Telescope ^21^, while gene expression counts for GENCODE v20 ‘PRI’ annotations were computed using RSEM v1.3.0.

Principal Component Analyses (PCA) were performed on retrotransposon and gene model counts, focusing on the top 1,000 most variable loci following variance-stabilizing transformation with DESeq2 v1.44.0 ^22^ within the R v4.4.2 statistical environment. Differential expression analyses were conducted using DESeq2, modeling disease state (Control or CD) and sex (Male or Female) as separate main effects to explain observed gene expression differences. Retrotransposons were considered differentially expressed if they exhibited a false discovery rate-adjusted p-value < 0.1 and an absolute fold-change > 0.5 for the disease state model term.

## Results

To assess the retrotransposon transcriptome in children with CD, clustering analyses were performed to determine the sample-to-sample distance using the CMU-ileal and rectal datasets. The retrotransposon transcriptome heatmap (**Figure 1A**) exhibited a clustering pattern similar to the gene transcriptome heatmap (**Figure 1B**). Specifically, ileal and rectal samples were grouped distinctly, and the CD samples were differentiated from non-IBD controls, allowing a clear distinction between the two groups.

**Figure 1.**
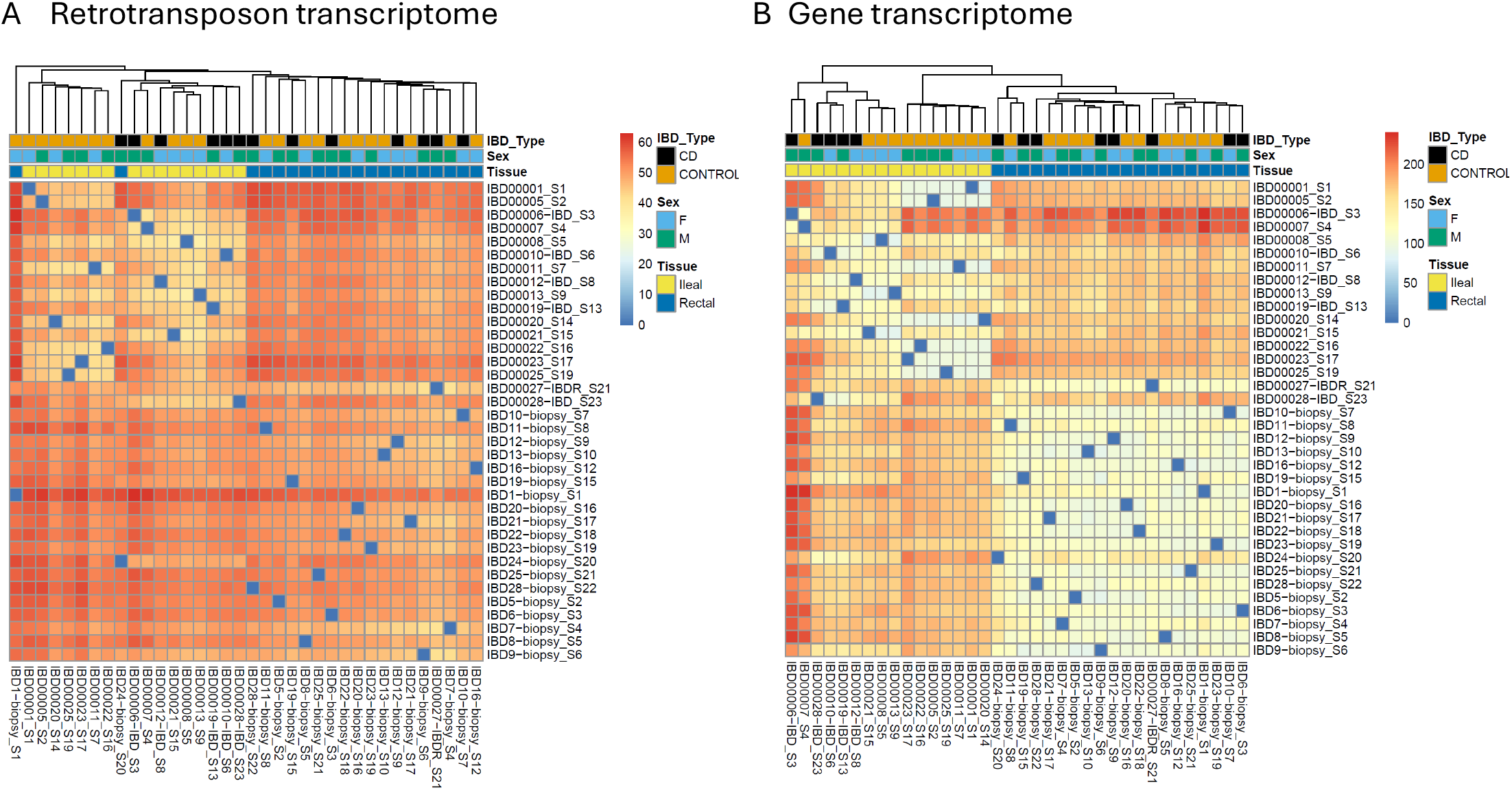
Sample-to-sample distance based on (A) retrotransposon transcriptome and (B) gene transcriptome

To gain further insights, an unsupervised PCA was conducted in both CMU-dataset and RISK dataset, based on gene and retrotransposon expression (**Figure 2**). In the CMU-ileal dataset, PC1 based on gene expression (**Figure 2A top**) and retrotransposon expression (**Figure 2A middle**) explained 56% and 37% of the variances, respectively, and broadly divided non-IBD controls to CD patients. This indicated that, similar to the gene transcriptome, the retrotransposon transcriptome is distinct in CD compared with non-IBD controls^14^.

**Figure 2.**
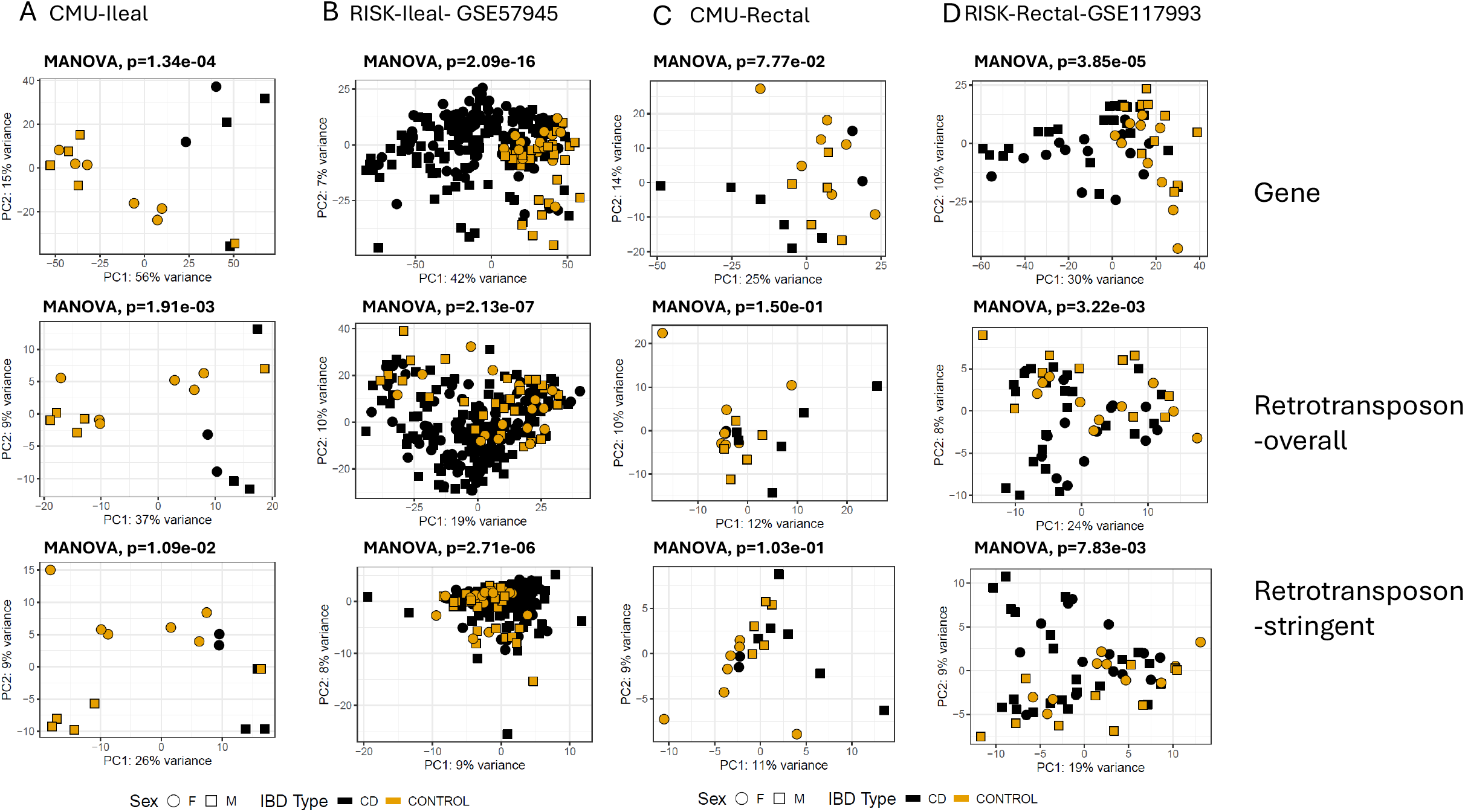
Principal component analysis (PCA) of sample similarity based on gene transcriptome and retrotransposon in ileal CMU and RISK cohort (A and B) and rectal CMU and RISK cohort (C and D)

To ensure that expression differences were not solely influenced by overlapping protein-coding genes, PCA was performed exclusively on retrotransposons with no positional overlap to known gene models (retrotransposon-stringent). In the public dataset, which was sequenced using single-end and unstranded short reads, PC1 derived from gene expression and overall retrotransposon expression explained 42% (**Figure 2B, top**) and 19% (**Figure 2B, middle**) of the variances, respectively. However, the separation based on retrotransposon-stringent is less distinct, accounting for only 9% (**Figure 2B, bottom**). The public dataset’s library preparation method hinders accurate mapping to repetitive regions, making it difficult to distinguish retrotransposon reads from overlapping genes on the opposite strand. Nevertheless, the PCA plot based on retrotransposon-stringent expression from the CMU-dataset, which was sequenced as paired-end and stranded with longer reads, still shows a clear separation between control and CD samples (26%, **Figure 2A, bottom**). This finding reinforces the conclusion that the expression patterns of retrotransposons are significantly different in ileal biopsies from CD patients compared to non-IBD controls.

In rectal biopsies, PC1 based on gene expression explained 30% and 25% of the variances in CMU-dataset and public dataset, respectively (**Figure 2C and D, top**). However, PC1 based on the overall retrotransposon transcriptome, showed less separation between the non-IBD controls and CD patients in both datasets (12% for CMU-rectal and 24% for RISK-rectal) (**Figure 2C and D, middle**). The separation was even less pronounced when considering PC1 based on the retrotransposon-stringent method (**Figure 2C and D, bottom**). Thus, subsequent analysis focused on ileal biopsy datasets.

Subsequently, differential expression analysis was conducted to pinpoint the differentially expressed retrotransposon loci (**Figure 3A** and **Supplemental Table 1**). In the CMU-ileal dataset, 118 retrotransposon loci with differential expression were identified, comprising 27 upregulated and 91 downregulated, specifically 74 belonging to the LINE-1 category and 44 to the HERV category. Notably, 15 retrotransposon loci were consistently downregulated across both datasets (CMU-ileal and GSE57945): HERV9N-int, HERV9-int (2 loci), HERVH-int (2 loci), HERVK22-int, LTR5A, HERVK-int, L1PA4 (5 loci), L1PA6, L1PA14 (**Figure 3B** and **Supplemental Table 2**). Eight of these 15 retrotransposon loci colocalize with 5 cellular genes (CUBN ^23^, EPS8L3 ^24,25^, CYP3A5, CYP2C18 ^26^, RPUSD3) (**Supplemental Table 2**). To further explore the relationship between retrotransposons and their adjacent genes, an analysis was performed using the Pearson correlation coefficient. The results (ranging from 0.64-0.94) indicated that 7 out of the 8 retrotransposons exhibited a positive linear correlation with their neighboring genes (**Supplemental Table 2**).

**Figure 3.**
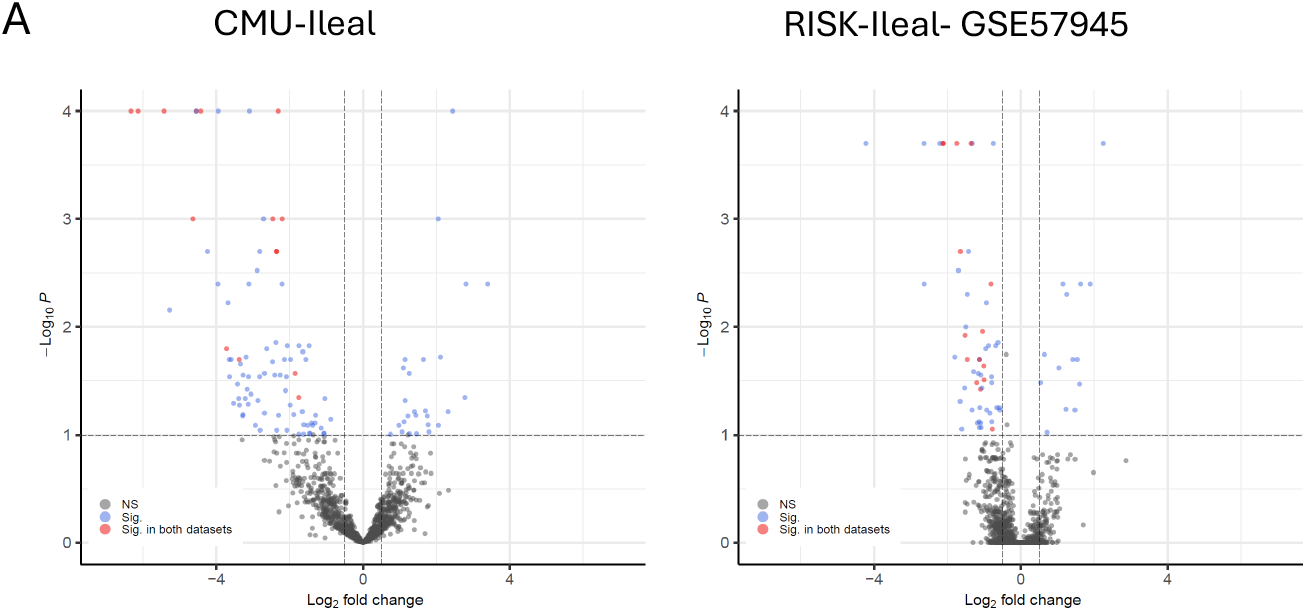
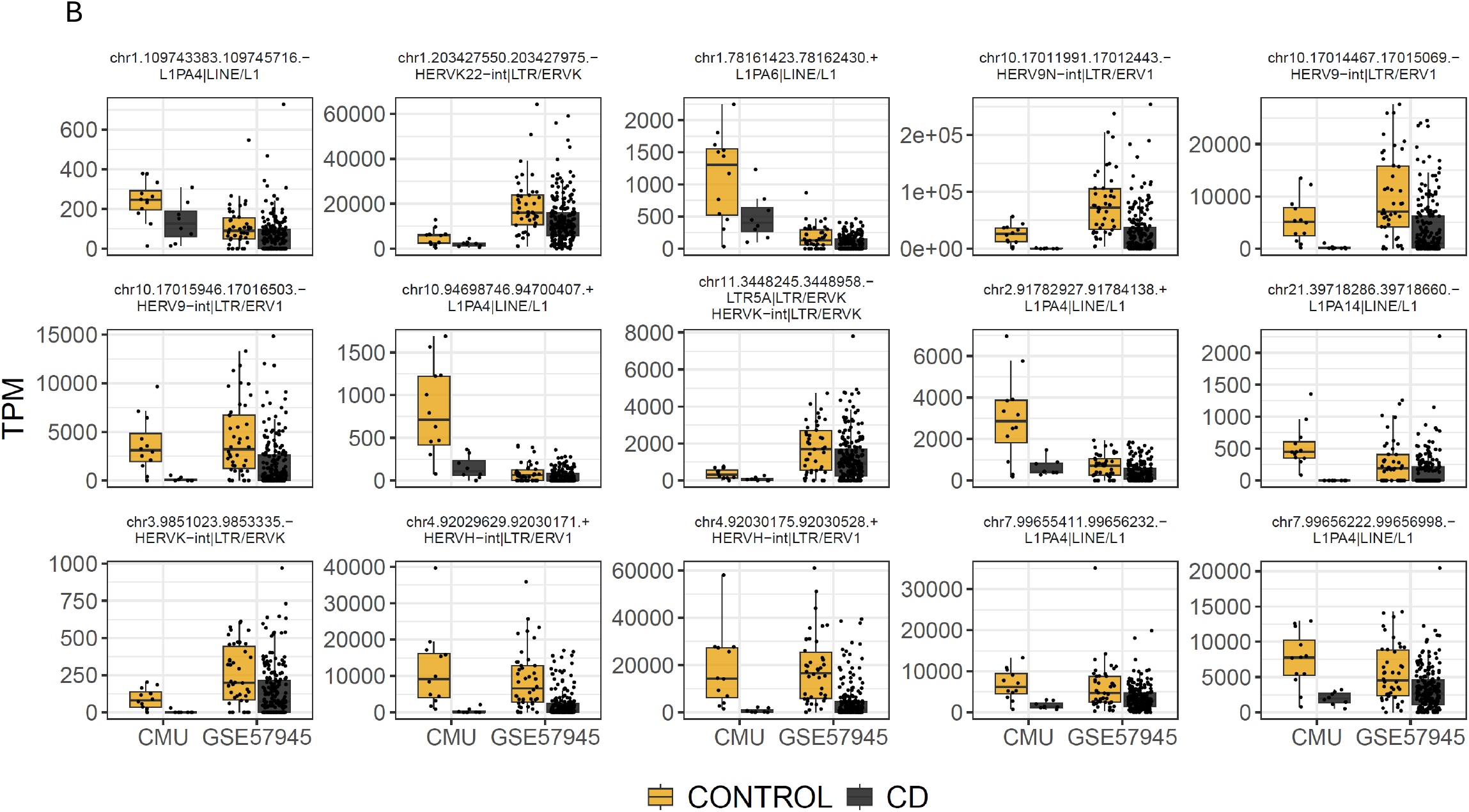
Differentially expressed retrotransposons in CD versus non-IBD controls. (A) Volcano plot of the differentially expressed retrotransposons. (B) Boxplots of the 15 downregulated retrotransposon loci common to both datasets.

## Discussion

For the first time to our knowledge, our study presents a detailed analysis of the retrotransposon transcriptome at the locus level in ileal and rectal biopsies from treatment-naïve children with CD and age-matched non-IBD controls. Children with CD exhibited a distinct pattern of retrotransposon transcriptome in ileal biopsies, with findings from our prospective cohort validated in a public dataset.

Retrotransposon transcription is precisely regulated across cell types and is tissue-specific, however, this regulation can be disrupted in inflammation ^10,11^. Our study found more downregulation of retrotransposon loci in children with CD. Although upregulation of retrotransposons can trigger immune responses and inflammation, other studies concur with our findings of more downregulation in inflammatory diseases ^5,6^. For example, a study investigating HERV transcriptome during SARS-CoV-2 infection, a known inflammatory response trigger, identified 210 HERV loci down-regulated compared to 72 loci upregulated ^27^. Furthermore, Klag et al. found most examined HERVs (including HERV-FRD, HERV-W, HERV-K-T47D, and HERV-KC4) were reduced in adult CD ileal biopsies ^10^.

Fifteen retrotransposon loci exhibited significant reductions across both datasets. Three cellular genes colocalized with the downregulated retrotransposon loci are linked to IBD. CUBN (HERV9, Chr10) demonstrated a decrease in histone methylation and expression in pediatric CD ^28^. EPS8L3 (L1PA4, Chr1) was associated with IBD in a genome-wide association study ^25^. Additionally, CYP2C18 (L1PA4, Chr10), involved in drug metabolism, is upregulated in colon biopsies from ulcerative colitis ^26^. Nevertheless, the precise impact of retrotransposons on neighboring genes and their independent functions remain unclear, necessitating further research given their involvement in inflammatory/autoimmune conditions.

Despite our novel findings, this study has several limitations. The small prospective cohort limited our ability to analyze retrotransposon correlations with disease characteristics. Furthermore, to avoid the potential interfering influence of treatment on retrotransposons, we assessed treatment-naïve children; however, this limits our understanding of retrotransposon involvement in treatment response. Our preliminary findings of downregulation of certain retrotransposons in CD warrant further mechanistic studies for a more comprehensive understanding of retrotransposons in CD.

## Author Contributions

*Concept and design*: Qing Chen, Suchitra K Hourigan. *Acquisition, analysis, or interpretation of data*: All authors. *Drafting of the manuscript*: Qing Chen. *Critical revision of the manuscript for important intellectual content*: All authors. *Statistical analysis*: Qing Chen, Colton McNinch. *Obtained funding*: Suchitra K Hourigan. *Supervision*: Suchitra K Hourigan. All authors reviewed and approved the final manuscript.

## Acknowledgements

The authors would like to acknowledge Justin Lack for his contribution to the ERVX pipeline.

## Data availability

Sequence data and biosample metadata have been deposited in the NCBI Sequence Read Archive (<u>PRJNA1251616</u>).

## Supplementary Tables

Table S1: List of differentially expressed retrotransposons in CD ileum

Table S2: List of 15 loci that downregulated across datasets

